# The evolution of anthropometric data quality methods and outcomes from 1993 to 2021 in the Comprehensive National Nutrition Survey and five rounds of the National Family Health Survey

**DOI:** 10.64898/2026.06.25.26356638

**Authors:** Pandara Purayil Vijin, Drishti Sharma, Robert Johnston, Phuong H. Nguyen, Sarang Pedgaonkar, S. K. Singh, Nadia Akseer, Purnima Menon, Zulfiqar A Bhutta, Rajesh Kumar

## Abstract

**Introduction:** High-quality anthropometric data is essential to creating effective nutrition policy, but there’s limited evidence on the quality of data from population-based household surveys in India, particularly at the sub-national level.

**Methods:** We compared survey procedures implemented by the National Family Health Survey (NFHS), rounds 1, 2, 3, 4, and 5, and the Comprehensive National Nutrition Survey (CNNS). We conducted a disaggregated analysis at the state level to observe quality performance for height-for-age (HAZ), weight-for-age (WAZ), and weight-for-height (WHZ) z-scores for children under five years. Nine data quality parameters, guided by WHO-UNICEF guidelines, were assessed: completeness of birth date and anthropometric measurement, sex ratio, age-heaping, position mismatch, digit preferences, implausible values, z-score distribution and growth faltering graphs.

**Results:** Survey methods improved over time, including enhanced training, increased use of computer-assisted devices, and real-time supervision. Consequently, from 1992 to 2020, there were improvements in anthropometric data quality, including more balanced age and sex distribution, increased completeness of birth date (from 53% to 95%) and anthropometric measures (from 60% to 91%), and decreased rounding of measurements. Steady albeit small improvements were observed in the standard deviation of HAZ (from 1.9 to 1.8) and WHZ (from 1.5 to 1.3). Implausible HAZ values were reduced from 5% to 2%. Data quality varied across states but generally showed a positive trend.

**Conclusions:** Anthropometric data quality in Indian population-level surveys shows satisfactory progress, reflecting global advancement. However, gaps persist, particularly in errors in birth date and height and weight measurement. Continued efforts are needed to further enhance data quality through improved survey planning, training, data collection, real-time supervision, analysis, and reporting, together with the strengthening of administrative data and civil records. High implausible values, especially in surveys with otherwise strong data quality, likely reflect, at least in part, underlying population inequities.

## Introduction

Achieving the UN’s Sustainable Development Goal 2 of “Zero Hunger” by 2030 will require significant progress in reducing childhood malnutrition [1], and reductions in global stunting and wasting are falling short. India, with high burdens of both stunting (35% in 2020) and wasting (19%), will require an accelerated rate of reduction to reach its goals [2,3]. Monitoring progress against malnutrition relies heavily on population-based surveys [4]. While studies suggest there has been a global improvement in anthropometric survey data quality over time, particularly with the application of standardized guidelines, data quality variations across countries and within India necessitate further investigation [5–7].

In India, the National Family Health Survey (NFHS) has been the chief source of child anthropometry estimates. The Ministry of Health also conducted the Comprehensive National Nutrition Survey (CNNS) in response to gaps in existing data in a number of areas, including micronutrient deficiencies in children [8–13]. Both surveys present data at the country and state levels, and both adhere to strict ethical guidelines [14]. NFHS has a broader scope than CNNS, which has a tighter focus on child nutrition. Their methodologies and target populations differ, making them complementary tools for policymakers.

However, there has also been confusion caused by significantly different data in surveys of similar geographic locations at close to the same time [15] The relatively slow progress against malnutrition despite government investments has added to this confusion.4 There have been post-hoc corrections and suggested alternative metrics, but these measures create their own concerns about data quality, especially at the sub-national level [7, 6, 17]

Addressing this gap, this study examines the trends in data quality for anthropometric measurements at the national and state level across NFHS (1993-2021) and CNNS (2016-18) surveys [4,8–13], identifying gaps and proposing improvements.

## Methods

### Data sources

We conducted a secondary analysis using publicly available data from NFHS rounds 1-5 (1992-3, 1998-99, 2005-2006, 2015-16, and 2019-21), and CNNS (2016-18). For NFHS data, we focused on the “child under age 5 born to a woman interviewed” recode files. For CNNS data, we analysed data for children under five years of age. Detailed methodological descriptions, including sample sizes for both surveys, are available elsewhere [4,8–13] (See S1 appendix Table 1-3).

### Comparative analysis of survey methodologies for data quality

To assess how data quality assurance practices have evolved over time, we conducted a comparative analysis of survey methodologies. This analysis examined a range of factors, including 1) the age groups of children included in the surveys, 2) the sampling techniques employed, 3) the sample sizes used, 4) the types of equipment utilized for data collection, 5) the training procedures for data collection personnel, 6) the standardization protocols in place to ensure consistency, 7) the methods used for collecting the data, and 8) the techniques employed for supervision and reporting of data quality.

### Anthropometric measures and analysis

We calculated z-scores for height-for-age (HAZ), weight-for-age (WAZ) and weight-for-height/length (WHZ), using WHO 2006 Child Growth Standards. Malnutrition prevalence estimates were survey-weighted to account for the sampling design. We employed the WHO STATA macro for analyzing anthropometric data [18]. Notably, non-weighted samples were used for all data quality indicators except sex ratio, following WHO guidelines [19].

### Data quality metrics

We assessed data quality using eight key indicators aligned with UNICEF and WHO metrics [19]. In addition to these metrics, we examined age-wise growth faltering as reflected in both national and state-level surveys [20, 21].

#### Birth date completeness

We assessed the proportion of children aged 0-59 months with complete recorded birth dates (day, month, and year) [19]. Per WHO-UNICEF guidelines, at least 90% of children are required to have complete birth dates for acceptable data quality [19, 22].

#### Missing anthropometric data

For data to be representative, completeness of height and weight information should be at 90%. We calculated completeness as the percentage of children aged 0-59 with both height and weight measurements. Cases of children without measurements (usually due to absence or refusal) were categorized as missing data [19, 22].

#### Sex ratio assessment

We assessed the sex ratio (males per 1000 females) among children aged 0-59 months. Sex ratios were calculated with sample weights for comparison to the reference population. Given the absence of a global cut-off, we compared the observed sex ratios to the expected ratios for this age group in India and individual states according to census data (1991, 2001, 2011) and projections for 2021. Chi-squared tests were employed to compare sex ratios from NFHS rounds 1-5 and CNNS with these reference values [19, 22].

#### Age heaping

We assessed age reporting quality to identify potential biases or misreporting. This can be indicated by age heaping (rounding up or down to preferred ages) or uneven age distributions across months and years. We employed two methods:1) visual inspection of the distribution of children by age in years and months, and 2) calculation of the absolute difference in mean HAZ between children born in January and December. Smaller differences suggest more accurate birth month reporting [19, 22, 23].

#### Measurement position mismatch

This section evaluates whether children were measured in the correct position according to their age. For children under 24 months, recumbent length is the recommended method, with standing height used for older children. Inaccurate positioning can lead to erroneous results. We assessed this using two methods:1) by calculating the proportion of children measured in the recommended position, and 2) creating line graphs to show the distribution of children measured by standing height, recumbent length, and those with missing measurements, categorized by age in months [19, 22].

#### Analysis of digit preferences

We examined potential inconsistencies in data collection and recording by analyzing digit preferences (a tendency to favour specific last digits when recording data, possibly indicating data fabrication or carelessness) in weight (0.1 kg) and height/length (mm) measurements. In addition to visualizing the distribution of terminal digits, we assessed digit preference using the Digit Preference Score (DPS) method.24 DPS considers both the extent and pattern of rounding errors. Higher DPS scores indicate a greater concentration of specific digits, suggesting potential problems. Generally, DPS values below 20 are considered acceptable [25].

#### Implausible z-scores

Z-scores (HAZ, WAZ, and WHZ) falling outside the WHO’s recommended ranges (HAZ: -6 to +6, WHZ: -5 to +5, WAZ: -6 to +5) are considered implausible. These values are typically a result of incorrect measurements, inaccurate recording of birth dates, or other data errors. A threshold of more than 1% implausible values suggests poor data quality.19 22 Note that “implausible” doesn’t mean “impossible,” and these could represent extreme forms of undernutrition.6 19 26 Several DHIS datasets exhibit a normal distribution of z-scores with tails extending beyond these ranges. Thus, the percentage of children below the cut-off also reflects children experiencing extreme z-scores.6 26 It is also important to note that anthropometric indicator calculations using the WHO macro automatically exclude out-of-range values (height <45CM and >110 cm for children < 24 months and <65 cm and >120 cm for children 24-59 months) [19, 27].

#### Z-score distribution analysis

As with implausible values, z-score distribution metrics can reflect real-world inequality, and are not solely indicators of quality [6, 19,26]. Nevertheless, evidence from multiple countries and surveys supports the use of SD as a quality indicator for anthropometric measures.28 We therefore analyzed the data quality for each state and survey by analyzing z-score distributions of HAZ, WAZ, and WHZ. We calculated the mean, standard deviations, and skewness/kurtosis. Deviations from normality were indicated by high standard deviations (HAZ>1.8 SD, WAZ>1.5 SD, and WHZ>1.8 SD), skewness (<-0.5 or >+0.5), and kurtosis (<2 or >4). 19 22 Kernel density plots provided a visual comparison of these distributions with the WHO MGRS for children aged 0-59 months [19, 29]. This comparison helped assess normality and identify potential data quality issues [28].

#### Growth faltering over time

We visualized age-specific z-scores using smoothed local polynomial regressions. Unexplained fluctuations (“waves” or “peaks”) in these graphs can indicate data quality problems [20,21].

### Data quality assessment

We assessed the data quality for 28 states and the Union Territory of Delhi using the indicators described earlier, comparing them to established benchmarks (See S2 Appendix Method 1). We employed visual inspection of distribution patterns, and calculated summary statistics (mean, range, standard deviation, interquartile range) to evaluate variability across states and surveys. Chi-square tests and student’s t-tests were used to compare data quality between surveys over time. All analyses were performed using Stata-SE version 17 [30].

## Results

### Variations in methodology and data quality

NFHS methodologies showed consistent improvements across rounds. While we did not identify major differences between NFHS and CNNS methodologies at a conceptual level, there could be variations in implementation that this study could only infer and not directly observe (Table 1).

**Table 1.** Summary of survey procedures to ensure data quality.

|  | NFHS-1 | NFHS-2 | NFHS-3 | NFHS-4 | NFHS-5 | CNNS |
| --- | --- | --- | --- | --- | --- | --- |
| Period over which surveys were conducted | Apr 1992 – Sept 1993 | Nov 1998 – Dec 1999 | Dec 2005 – Aug 2006 | Jan 2015 – Dec 2016 | Jun 2019 - May 2021 | Feb 2016 – Nov 2018 |
| Survey Duration | 1 year and 4 months | 1 year and 1 month | 9 months | 1 year and 10 months | 1 year 11 Months | 2 years and 8 months |
| Number of phases data collection | One | Two | Two | Two | Two | Three |
| Estimates at state/district level | State | State | State | District and State | District and State | State |
| Number of states and Union territories (UT) covered | 24 states & 1UT | 25 states & 1UT | 28 states & 1UT | 29 states & 7 UTs | 28 states & 8 UTs | 29 states & 1 UT |
| Age (months) covered for anthropometry | 0-47 | 0-35 | 0-59 | 0-59 | 0-59 | 0-59 |
| Age estimation method | Century Month Codes (CMC) | Century Month Codes (CMC) | Century Month Codes (CMC) | Century Month Codes (CMC) | Century Day Code (CDC) | Century Day Code (CDC) |
| Mode of survey | Paper-and-Pencil | Paper-and-Pencil | CAPI with paper-based collection of anthro data |  |  |  |
| Number of questions in the survey questionnaire | 368 | 344 | 756 | 981 | 1242 | 384 |
| SAMPLING |  |  |  |  |  |  |
| Sample size | 48,959 | 33,026 | 51,555 | 2,59,627 | 232,920 | 38,060 |
| Sampling design | Multi-stage sampling design with probability proportional to population size (PPS) after stratification was done by district, separated into rural and urban areas. |  |  |  |  |  |
| Sample is sufficiently powered to provide malnutrition estimate at state level | Yes | Yes | Yes | Yes | Yes | Yes |
| EQUIPMENT |  |  |  |  |  |  |
| Equipment met WHO/UNICEF specifications | Yes | Yes | Yes | Yes | Yes | Yes |
| Weighing scale | Salter Scale | Digital SECA Scale |  |  |  |  |
| Height/length | Height measuring wooden board |  |  | Length: SECA 417 Infantometer (0-23 months)<br>Height: SECA 213 Stadiometer (24-59 months) |  | Three-piece wooden board |
| TEAM COMPOSITION |  |  |  |  |  |  |
| Number of Health Investigators (HI) | No separate HI | 1HI/ Team | 2HI/Team | 2HI/Team | 2HI/Team | 2HI/ Team |
| Number of anthropometrist in team | 1 | 1 | 2 | 2 | 2 | 2 |
| Minimum education qualification | No | Yes | Yes | Yes | Yes | Yes |
| **TRAINING OF FIELD WORKERS** |  |  |  |  |  |  |
| Duration of comprehensive training | 3 weeks | 3 weeks | 3.5 to 4 weeks | 4 Weeks | 4 Weeks | 4 Weeks |
| Training Method | Centralized | Centralized | Centralized | Cascade | Cascade | Cascade |
| **ANTHROPOMETRY STANDARDIZATION** |  |  |  |  |  |  |
| Standardisation | No | No | No | Yes | Yes | Yes |
| Reporting of Standardisation | No | No | No | No | No | Yes |
| **SUPERVISION** |  |  |  |  |  |  |
| Technical supervision | 3-tier monitoring | 3-tier monitoring | 3-tier monitoring | 3-tier monitoring | 3-tier monitoring | 3-tier monitoring |
| **QUALITY ASSURANCE** |  |  |  |  |  |  |
| Re-measurements | No | No | No | No | No | Yes<sup>1</sup> |
| Field check Tables | No | No | No | Yes | Yes+ POQR<sup>2</sup> | Yes |
| **DISSEMINATION** |  |  |  |  |  |  |
| Reporting on the quality of the anthropometric data is included in final reports | No | No | No | No | No | Yes |
| Public availability of survey data | De-identified survey data publicly available |  |  |  |  | Govt endorses data, authorises release to researchers |

<sup>1</sup>In CNNS, height was measure twice whereas weight was measured once.
<sup>2</sup>NFHS-5 developed and used an algorithm called the Project Officer's Query Report (POQR) on the Supervisor's CAPI instrument.

#### Survey design and sampling

NFHS surveys were conducted every 5-10 years. There was some overlap in data collection periods between CNNS and NFHS-4 in certain states. Both surveys employed multi-stage, stratified sampling methods. Earlier NFHS surveys were designed to report anthropometric indicators at the state level, allowing for smaller sample sizes. However, NFHS-4 and 5 aimed for district-level representation, necessitating larger samples.

#### Age of children

Children under 48 months were included in NFHS-1, while NFHS-2 limited the age range to under 36 months. Subsequent surveys in both NFHS and CNNS expanded the focus to children under five years old (0-59 months).

#### Personnel

More resources were devoted to anthropometric measurements over time. In NFHS-1, general field investigators handled measurements, while NFHS-2 introduced a dedicated health investigator for measurements, and by NFHS-3, there were two qualified health investigators assigned to this task, as there were in CNNS.

#### Training approach

Training methods shifted from centralized programs (NFHS-1 to 3) to a cascade model (NFHS-4, 5 and CNNS), which involves international experts training trainers, who then train local trainers. This ensured a consistent flow of well-trained investigators across locations.

#### Questionnaires and data capture

The number of questions per survey increased from 368 to 1242 between NFHS-1 and NFHS-5. CNNS had a relatively shorter questionnaire, with 384 questions. The survey questionnaires transitioned from paper to Computer Assisted Personal Interviews (CAPI) after 2005.

#### Measurement equipment and standardization

All surveys used recommended SECA-brand digital scales for weight measurements. Height measurement equipment varied: NFHS-1 to 3 and CNNS employed the gold standard Shorr height boards, while NFHS-4 and 5 adopted SECA infantometers and stadiometers. Regular calibration with standard weights and rods ensured accuracy in NFHS-4, 5, and CNNS. Notably, CNNS publicly reported their standardization findings, while those for NFHS surveys were not made public [13]. Lastly, CNNS employed an additional layer of quality control by taking all anthropometric measurements (except weight) twice [31].

#### Supervision and monitoring

All NFHS and CNNS surveys ensured data quality through a three-tier monitoring system with technical supervision. This system involved surveys being conducted by team supervisors, field coordinators, resource personnel from field agencies, and key investigators from associated organizations. Enhanced monitoring in NFHS-4, 5, and CNNS included digitalised field check tables for real-time tracking and feedback of data completeness, age heaping, and digit preference.

### Data quality parameters

Characteristics of the anthropometric data quality indicators across survey rounds at the national and state levels are summarized in Table 2 (See S3 appendix Table 4).

**Table 2.**
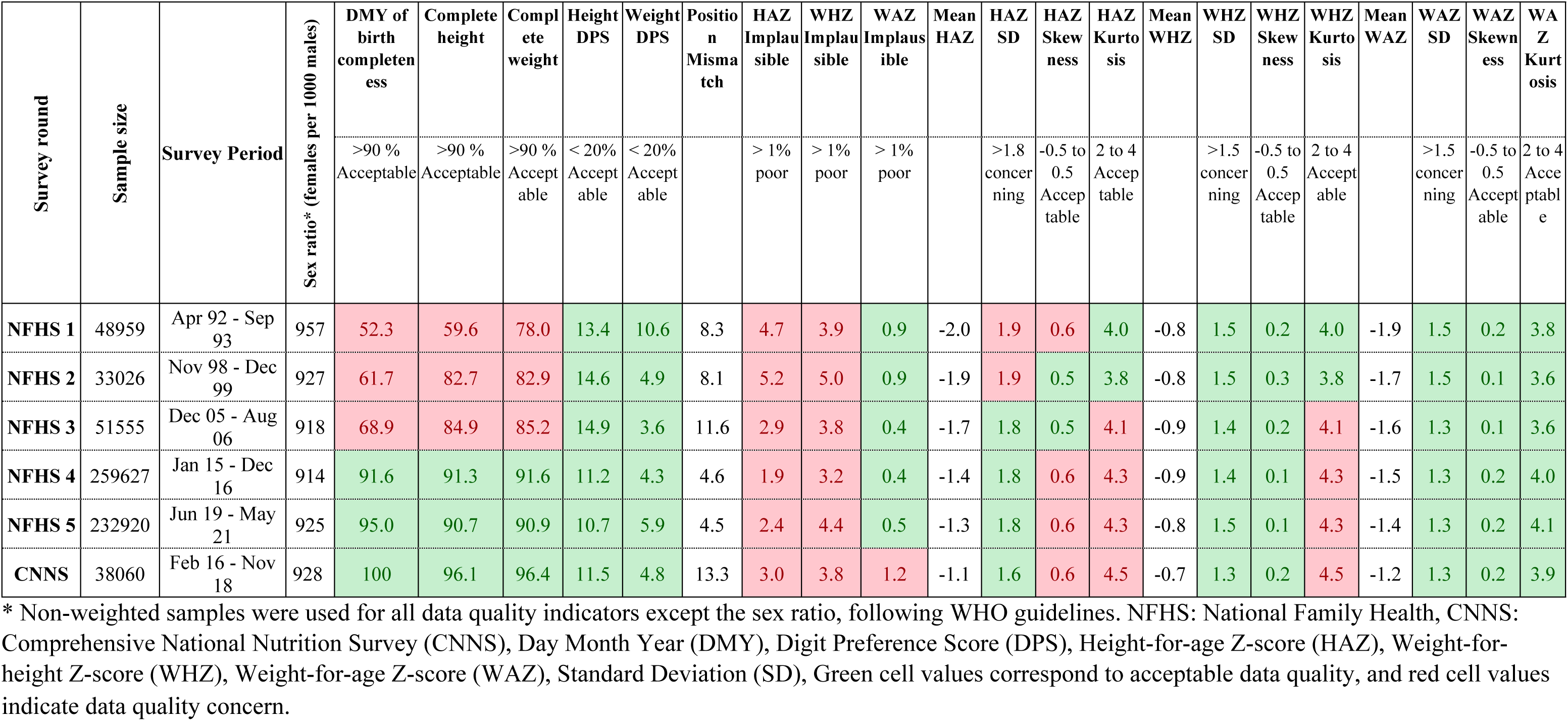
Summary statistics of anthropometric data quality indicators for India for NFHS1-5 and CNNS.

| Survey round | Sample size | Survey Period | Sex ratio* (females per 1000 males) | DMY of birth completeness | Complete height | Complete weight | Height DPS | Weight DPS | Position Mismatch | HAZ Implausible | WHZ Implausible | WAZ Implausible | Mean HAZ | HAZ SD | HAZ Skewness | HAZ Kurtosis | Mean WHZ | WHZ SD | WHZ Skewness | WHZ Kurtosis | Mean WAZ | WAZ SD | WAZ Skewness | WAZ Kurtosis |
| --- | --- | --- | --- | --- | --- | --- | --- | --- | --- | --- | --- | --- | --- | --- | --- | --- | --- | --- | --- | --- | --- | --- | --- | --- |
|  |  |  |  | >90 % Acceptable | >90 % Acceptable | >90 % Acceptable | < 20% Acceptable | < 20% Acceptable |  | > 1% poor | > 1% poor | > 1% poor |  | >1.8 concerning | -0.5 to 0.5 Acceptable | 2 to 4 Acceptable |  | >1.5 concerning | -0.5 to 0.5 Acceptable | 2 to 4 Acceptable |  | >1.5 concerning | -0.5 to 0.5 Acceptable | 2 to 4 Acceptable |
| NFHS 1 | 48959 | Apr 92 - Sep 93 | 957 | 52.3 | 59.6 | 78.0 | 13.4 | 10.6 | 8.3 | 4.7 | 3.9 | 0.9 | -2.0 | 1.9 | 0.6 | 4.0 | -0.8 | 1.5 | 0.2 | 4.0 | -1.9 | 1.5 | 0.2 | 3.8 |
| NFHS 2 | 33026 | Nov 98 - Dec 99 | 927 | 61.7 | 82.7 | 82.9 | 14.6 | 4.9 | 8.1 | 5.2 | 5.0 | 0.9 | -1.9 | 1.9 | 0.5 | 3.8 | -0.8 | 1.5 | 0.3 | 3.8 | -1.7 | 1.5 | 0.1 | 3.6 |
| NFHS 3 | 51555 | Dec 05 - Aug 06 | 918 | 68.9 | 84.9 | 85.2 | 14.9 | 3.6 | 11.6 | 2.9 | 3.8 | 0.4 | -1.7 | 1.8 | 0.5 | 4.1 | -0.9 | 1.4 | 0.2 | 4.1 | -1.6 | 1.3 | 0.1 | 3.6 |
| NFHS 4 | 259627 | Jan 15 - Dec 16 | 914 | 91.6 | 91.3 | 91.6 | 11.2 | 4.3 | 4.6 | 1.9 | 3.2 | 0.4 | -1.4 | 1.8 | 0.6 | 4.3 | -0.9 | 1.4 | 0.1 | 4.3 | -1.5 | 1.3 | 0.2 | 4.0 |
| NFHS 5 | 232920 | Jun 19 - May 21 | 925 | 95.0 | 90.7 | 90.9 | 10.7 | 5.9 | 4.5 | 2.4 | 4.4 | 0.5 | -1.3 | 1.8 | 0.6 | 4.3 | -0.8 | 1.5 | 0.1 | 4.3 | -1.4 | 1.3 | 0.2 | 4.1 |
| CNNS | 38060 | Feb 16 - Nov 18 | 928 | 100 | 96.1 | 96.4 | 11.5 | 4.8 | 13.3 | 3.0 | 3.8 | 1.2 | -1.1 | 1.6 | 0.6 | 4.5 | -0.7 | 1.3 | 0.2 | 4.5 | -1.2 | 1.3 | 0.2 | 3.9 |

\* Non-weighted samples were used for all data quality indicators except the sex ratio, following WHO guidelines. NFHS: National Family Health, CNNS: Comprehensive National Nutrition Survey (CNNS), Day Month Year (DMY), Digit Preference Score (DPS), Height-for-age Z-score (HAZ), Weight-for-height Z-score (WHZ), Weight-for-age Z-score (WAZ), Standard Deviation (SD), Green cell values correspond to acceptable data quality, and red cell values indicate data quality concern.

#### Sample representativeness and data completeness

NFHS and CNNS surveys achieved close-to-expected sex ratios based on census data (See S4 appendix Table 5). There were significant improvements in date-of-birth (DOB) completeness over time, from 54% in NFHS-1 to 95% in NFHS-5 (Table 2). Most states reached acceptable DOB completion (>90%) by NFHS-4. CNNS had exceptional DOB completeness: 100% across all states but one (See S3 Appendix 3 Table 4 and S5 appendix 5 Fig 1).

Visual inspection of age distributions (by years, months, and specific birth month) suggests reduced age heaping across NFHS surveys. NFHS-4 and 5 showed the most uniform age distributions across states and rounds, while CNNS heaping resembled NFHS-3. Detailed state-wise distributions are available in S6 appendix Fig 2-19.

The completeness of height and weight measurements in NFHS surveys increased significantly between NFHS-1 and 4, rising from 60% to 91% for height and 78% to 91% for weight. An increasing number of states met the criteria for complete height and weight data. However, this trend reversed in NFHS-5, in which ten out of 28 states reported higher proportions of missing anthropometry data than in NFHS-4 (7 out of 28). On the other hand, CNNS surveys achieved higher completeness than NFHS surveys, with all states exceeding 90% except West Bengal (82%) (See S7 Appendix Fig 20-21).

#### Measurement errors in age estimation

As with age distribution, distribution by birth month should be uniform if measurements are accurate.23 We observed variations across states and surveys, with unexpected peaks in specific months (See S6 Appendix Fig 14-19). The January-December difference in mean HAZ suggested potential errors in reported birth month, with higher errors in NFHS compared to CNNS. Notably, from NFHS-1 to NFHS-3 states with significant difference between mean HAZ between January and December decreased and rose again for NFHS-4 (20 out of 29) and NFHS-5 (21 out of 29) (See S8 Appendix 8 Table 6).

To visualize these trends, we plotted HAZ by month of birth across all rounds (See S8 Appendix Figure 22) [23]. NFHS-1 and 2 showed a slight decline in HAZ from January to June, followed by a sharp rise from July to December. This pattern was less pronounced in later rounds. CNNS, however, lacked this trend, suggesting more accurate birth month reporting. Additionally, analysis of HAZ by months within a completed year (0-11) revealed a subtler pattern of age estimation bias, one opposite to the birth month trend (S8 Appendix Fig 23)

### Measurement errors in height and weight: positioning and digit preference

Improper positioning during height measurements occurred in some cases. Across states, the proportion of children measured in the wrong position ranged from 2%-20% for NFHS-1, 0.1%-20% for NFHS-2, 0.5%-19% for NFHS-3, 1%-9% for NFHS-4, 2% to 7% for NFHS-5 and 2%-20% for CNNS (Fig1). Clearly, these errors decreased over time in NFHS surveys, with NFHS-5 showing the lowest range (1% to 7%). CNNS had a wider range of positioning errors (5% to 20%) compared to NFHS-4 and NFHS-5 (S3 Appendix Table 4 & S9 Appendix Figure 24-29).

**Fig 1.**
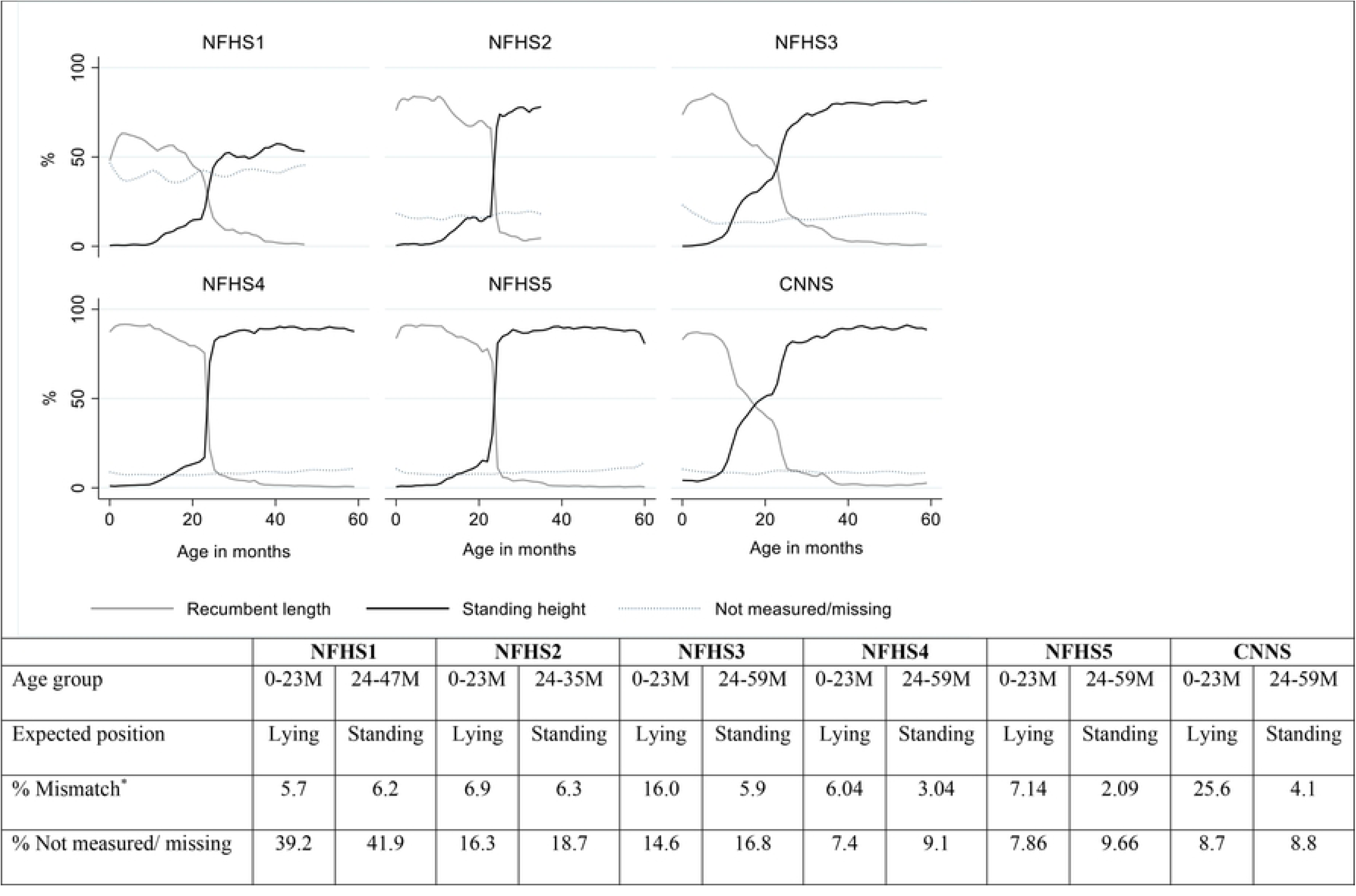
Proportions of anthropometric measurements with mismatched positioning (length/height versus recommended guidelines) for India across NFHS 1-5 and CNNS. Note: *Mismatch means children under 24 months were measured standing (height) or children 24 months or older were measured laying down (recumbent length), as opposed to the recommendation. NFHS: National Family Health Surveys. CNNS: Comprehensive National Nutrition Survey

NFHS-3 through 5 showed acceptable digit preference scores (DPS) for height (below 20) for India (Table 2). However, some states exhibited high DPS in earlier rounds, particularly NFHS-1 and NFHS-2 (e.g., Nagaland and Arunachal Pradesh). The quality particularly improved in NFHS-4 and 5, with all states achieving acceptable DPS for height (S3 Appendix 3 Table 4 & S10 Appendix Fig 30-35).

For weight, most states maintained acceptable DPS for all rounds. Erroneously rounding to zero or five was the most common issue, decreasing over time. The average error score dropped from 11 in NFHS-1 to 3.6 in NFHS-3 but increased to 6 in NFHS-5 (S3 Appendix Table 4 & S10 Appendix Fig 36-41).

#### Visualizing z-score distributions

We compared the distribution of HAZ, WAZ, and WHZ for children under 5 using kernel density across different surveys (CNNS and the last three rounds of NFHS) with the WHO reference population. Fig 2 includes HAZ/WAZ/WHZ distributions for India and select states compared to the WHO Growth Standard. Online supplemental appendix 11 Fig 42-44 provides detailed plots for each state across surveys.

**Fig 2.**
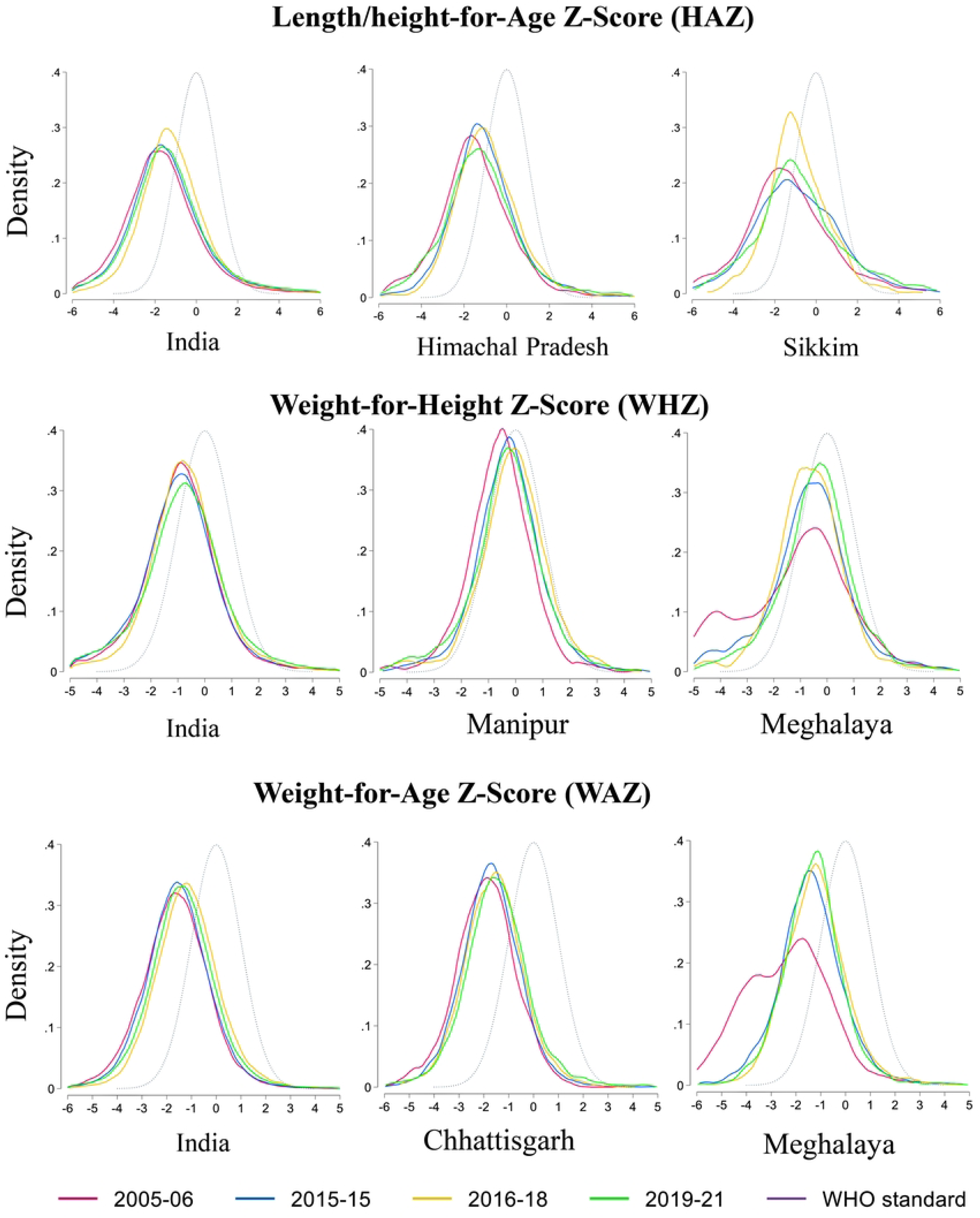
Kernel density plots of Height-for-Age, Weight-for-Height, and Weight-for-Age Z-score distributions in India and selected states Note: NFHS: National Family Health Surveys. CNNS: Comprehensive National Nutrition Survey. WHO: World Health Organization

The quality of the distributions was assessed based on smoothness and conformity to a bell-shaped curve (absence of bimodality or skewness). The z-scores were near-normally distributed. When compared with the WHO standard, these showed a leftward shift of the entire distribution. While most states transitioned to smoother curves in the distribution of z-scores over time, a few (e.g., Arunachal Pradesh, Meghalaya, Nagaland, and Sikkim) showed deviations from this pattern. Extreme z-scores, such as -6 SD for HAZ or -5 SD for WHZ, appeared as part of the continuous distribution rather than as exceptional outliers.

Similarly, visualizing child growth faltering curves across age reveals typical patterns for India and its states for HAZ, WHZ, and WAZ in children under 5. All three indicators start below zero at birth. WHZ remains relatively constant after birth, while HAZ and WAZ typically show a dip between 2-6 months and 12-18 months before stabilizing. Mean HAZ at birth increased with time across surveys, except for NFHS-5, which showed an unusually lower mean HAZ at birth compared to the rest of the surveys (S11 appendix Fig 45-47) [32].

#### Dispersion in height-related measures (HAZ and WHZ)

Height-related measures (HAZ, WHZ) showed more data quality issues than the weight-related measure (WAZ). Height-for-age (HAZ) distributions were flatter (kurtosis>4) and more positively skewed (i.e., with a tail extending to the right, skewness>+0.5) than WHZ and WAZ curves at the national level in all surveys. While some states in NFHS-3 and CNNS showed skewness within the acceptable limit (<0.5), the national trend was slightly skewed (S3 Appendix Table 4 & S12 Appendix 12 Fig 48). In comparison, weight-for-height (WHZ) distributions for India and most states fell within the acceptable range (-0.5 to 0.5). However, similar to HAZ, kurtosis in WHZ often exceeded the recommended limit (2-4) in most states.

Next, we examined standard deviation (SD). The median standard deviations of HAZ and range of SD (Min - Max) across states for NFHS-1 to 5 and CNNS were 1.8 (1.6 - 2.0), 1.8 (1.7 - 1.9), 1.7 (1.6-1.9), 1.7 (1.7 - 1.9), 1.8 (1.7-1.9) and 1.5 (1.5 -1.6), respectively (Fig 3 and S3 Appendix Table 4). CNNS achieved a remarkably lower median SD (1.5). High SD (>1.8) indicates potential data quality issues. Nearly half of the states in NFHS rounds 1 and 2 had high SD (>1.8), improving to one-quarter by round 3. However, this improvement didn’t continue. The proportion with high SD increased again in round 4 and worsened further in round 5, with half the states exceeding the cut-off once more. In contrast, only three out of 28 states (10%) in CNNS had high SD (S3 Appendix Table 4 & S12 Appendix Fig 48).

**Fig 3.**
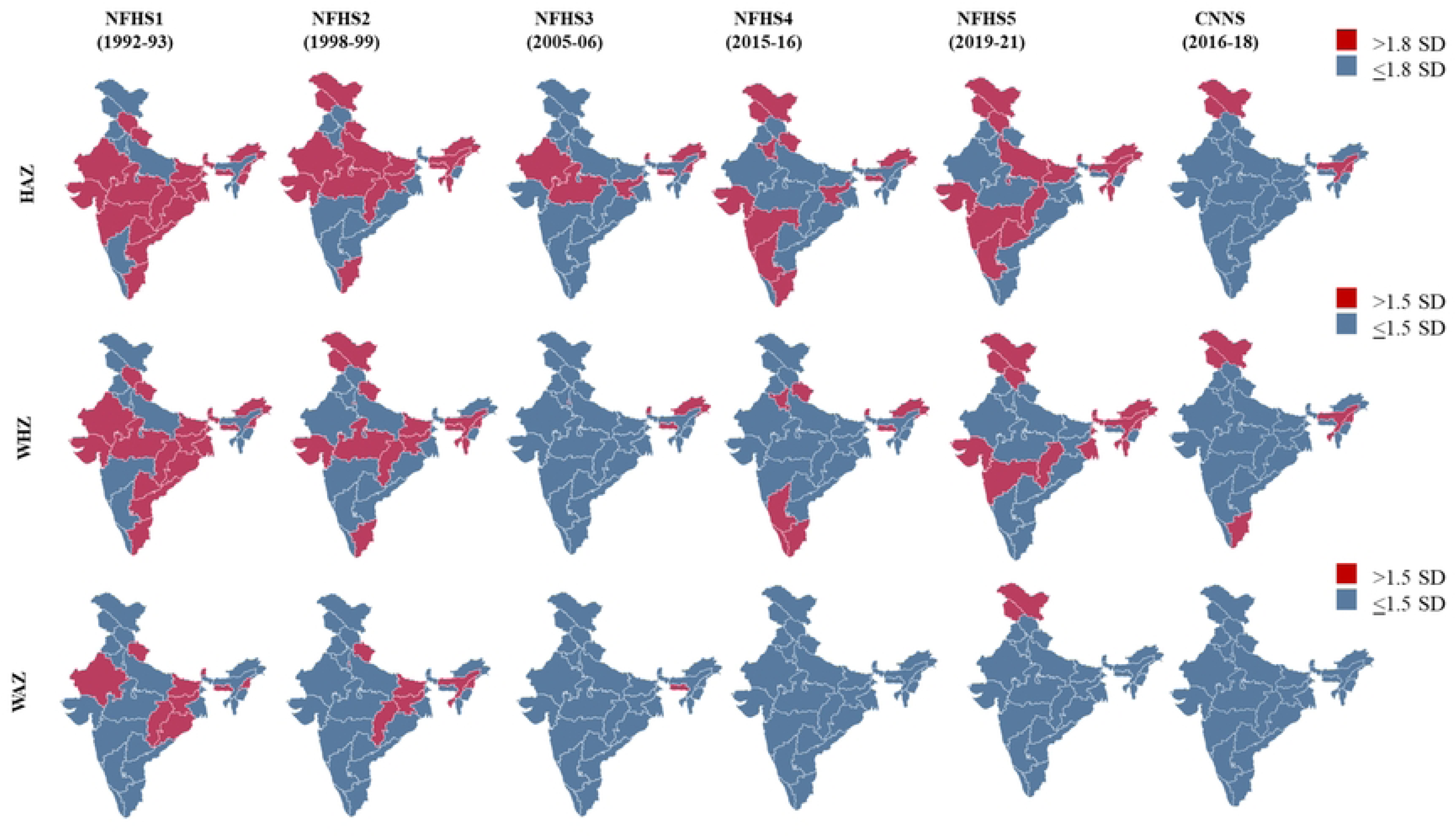
Subnational variations in standard deviation of Height-for-Age, Weight-for-Height, and Weight-for-Age Z-scores in India reported across NFHS 1-5 and CNNS Note: Standard deviations (SDs) cut-offs used in the above maps have been referred from Technical Notes from the background document for country consultations on the 2021 edition of the UNICEF-WHO-World Bank Joint Malnutrition Estimates, 2021. HAZ: Height-for-Age Z-score, WHZ: Weight-for-Height Z-score, and WAZ: Weight-for-Age Z- score. NFHS: National Family Health Surveys. CNNS: Comprehensive National Nutrition

High SD (>1.5) suggests potential data quality issues. A limited number of states fell into this category across all surveys, with the highest proportion in NFHS-1 (7 out of 19 states) and the lowest in CNNS (5 out of 28) (S3 Appendix Table 4 & S12 Appendix Fig 48).

Despite the distributions being normal (as indicated by SDs and skewness values), all surveys exceeded the acceptable limit (1%) for implausible height-for-age (HAZ) and weight-for-height (WHZ), both in India and across most states (Table 2).

However, the proportion of flagged values decreased from NFHS-1 to NFHS-4, with NFHS-4 showing the lowest overall proportion of implausible z-scores. Even in NFHS-4, the majority of states exceeded the 1% acceptable limit. For NFHS-4, flagged HAZ values ranged from 0.6% to 3.6%, and flagged WHZ values ranged from 0.8% to 5.7% (S3 Appendix Table 4 & S13 Appendix table 7).

#### Dispersion in weight for age z-score (WAZ)

Standard deviations (SD) of weight-for-age (WAZ) showed a slight improvement across NFHS rounds, ranging from 1.2 to 1.3 (S3 Appendix Table 4 & S12 Appendix Fig 48). Notably, all states in NFHS-4 or later had an SD within the 1.5 limit, indicative of good data quality. In contrast, a few states in earlier rounds (five in NFHS-1) fell into this category. Overall, skewness of WAZ distributions remained within the acceptable range (-0.5 to 0.5) for both India and state levels. However, similar to HAZ and WHZ, kurtosis often exceeded the recommended upper limit (>4) in about half of the states (S3 Appendix Table 4 & S12 Appendix Fig 48).

Weight-for-age (WAZ) z-scores had the lowest proportion of implausible values compared to HAZ and WHZ. Across all NFHS rounds and CNNS, flagged values remained consistently below 1.2%, including state-level data (See S3 Appendix Table 4 & S13 Appendix Table 6).

#### Regional variation in data quality across time

Clearly, the number of states across the country meeting the data quality criteria of completeness increases through rounds 1-5 of NFHS. The only exception in the trend is NFHS-5, in which a greater number of states could not meet completeness criteria for height and weight measurements. Errors in measuring height and weight reduced across time. Notably, for NFHS-4 and 5 all states met the quality criteria for positioning as well as rounding errors in weight and height (S14 Appendix Fig 49-51). The difference between mean HAZ from January to December reduced from NFHS-1 to NFHS-3, but a greater number of states reported significant difference in NFHS-4 and 5 (S8 Appendix Table 5). Almost all states met the WAZ SD criteria in NFHS-3, 4 and 5. Similarly, a greater number of states meet SD criteria for HAZ and WHZ in NFHS-3 than in NFHS-4 and 5 (Fig 3). Despite this improving trend in SD of HAZ and WHZ, almost all states fell short of the criteria for biologically implausible values, even in the survey rounds that demonstrated minimum measurement errors.

Andhra Pradesh, and Karnataka consistently lagged in completeness of height and weight. In the case of DOB completeness Uttar Pradesh and Madhya Pradesh fell short of the criteria for most of the surveys except NFHS-5 and CNNS (appendix p55). States that showed a reduced difference between January and December HAZ in more recent rounds were West Bengal, Tamil Nadu, Himachal Pradesh, Mizoram, Sikkim, and Tripura. For the last two rounds, Karnataka, Maharashtra, Gujrat J&K, Arunachal Pradesh, and Sikkim consistently fell short of criteria for the SDs in HAZ and WHZ (S14 Appendix Fig 52-60).

## Discussion

This study assessed anthropometric data quality at the state level in all rounds of India’s NFHS and CNNS surveys, following WHO/UNICEF guidelines. We found significant national and state-level improvements, including better age and sex distribution, complete birthdate records, accurate height/weight measurements, reduced measurement position errors, and less rounding. The improvements in quality likely stem from evolving survey protocols, as well as systemic changes in health access and civil records. The differences between states can be best explained by variations in implementation. The workload in the recent NFHS surveys and disruption during COVID-19 can likely explain some data quality properties of recent NFHS surveys. Even with the improvements, there’s room for a further reduction in measurement errors.

### Improved NFHS procedures lead to more complete measurements and fewer errors

The NFHS has improved procedures for better data quality over the years. This is evident in better-trained personnel, investments in state-of-the-art equipment, validation exercises, and the utilization of technology, particularly in monitoring data quality through field check tables.33 CNNS’s procedures matched those of NFHS [13].

These improved procedures are clearly reflected in the eventual achievement of near-complete data collection for age, height, and weight; reduced measurement errors such as position mismatch and digit preference; the reduction of out-of-range height and weight values; and smaller standard deviations.

Our analysis supports the common view that measuring height (i.e., Height-for-Age (HAZ) data, and Weight-for-Height/Length (WHZ)) is more difficult and prone to error than weight (Weight-for-Age (WAZ)), especially among children under two [34]. As reported elsewhere, anthropometric data quality is poorer for children under two years of age, as demonstrated by the higher standard deviation of z-scores [28, 35]. The proportion of flagged data is higher in younger children in NFHS-4 [7]. Measurement error in this age group has a greater potential to derange the z-score calculation, as even 5mm or 100g of variation can cause shifts, leading to an incorrect assessment of nutritional status. Anthropometry trainings and real-time monitoring have likely been instrumental in reducing errors for this age group in more recent surveys.

We clearly observed state-level differences in data quality parameters, which have been previously attributed to sociodemographic, geographical, and implementation variations, especially team-related factors, such as the type of agency doing the work and each team’s work load [7, 34]. Nevertheless, it is worth noting that even states with the most difficult terrains, such as in Manipur and Mizoram, have shown satisfactory data quality across all indicators, demonstrating that good quality is achievable despite constraints.

In addition to methodological changes, we should also consider how the overall health and data ecosystem of a region can explain some of the observed patterns; specifically, how higher birth registration rates likely contributed to improved survey data quality [36]. National birth registration coverage increased from 55% in 2001 to 79% in 2015 due to the nationwide strengthening of the Civil Registration System, and states with higher birth registration completeness also report more complete birth records in surveys [36–38]. Increased birth registration might also be linked to better access to maternal healthcare and more institutional deliveries [10–12].

### Insights from NFHS and CNNS comparisons

CNNS had a smaller scope and sample size compared to NFHS-4 and 5, and showed somewhat higher data quality, likely due to implementation rather than methodology. Its stronger emphasis on training, reporting completeness, and more accurate birth date estimation may explain its nearly complete age reporting and reduced January-December HAZ differential [17]. This more complete data, coupled with improved age estimation, is likely responsible for its relatively tighter HAZ and WHZ standard deviations. CNNS’s reporting of data quality estimates also made it more transparent, which also likely contributed to quality [34].

However, the prevalence of implausible values was higher in CNNS than in NFHS, despite CNNS’s three-tiered supervision and implementation of Gibson’s double measurement protocol (with a third measurement if the initial values differed by ≥5mm or ≥100g). This suggests that for large-scale surveys like NFHS and CNNS, the efficacy of the Gibson method may be limited due to its potential for redundant measurements, without a proportional improvement in data quality. Furthermore, a focus on real-time data collection and immediate feedback from a dedicated team, a practice increasingly adopted in the NFHS from round 4 onwards, might be a more efficient approach than multi-tiered monitoring.

A larger survey size and longer questionnaires could have contributed to higher errors and thus higher standard deviation in HAZ for NFHS-4 and 5 as compared to CNNS [7,34,39]. NFHS-5 also introduced a key methodological change that might have created confusion: Age reporting shifted from the Century Month Code to the Century Day Code[40]. While secular trends suggest there should have been continuous improvement in HAZ score across all ages, lower reported at-birth HAZ scores in NFHS-5 might have resulted from this change in methodology, and warrants further investigation [32].

The two-phase design of NFHS-5, with one phase occurring during the COVID-19 pandemic, also likely contributed to data quality challenges. There were more missing height measurements (nearly 10% for children under five) and wider standard deviations in NFHS-5 than in NFHS-4, at both the national and state level [41]. COVID-19 restrictions might have led to strained supervisory and team capacity and contributed to higher refusal and error rates [34].

### Are the flagged z-score values implausible?

The challenge of implausible values and standard deviations of z-scores within the data is rooted in both measurement errors and inequities within the population [6, 26]. Undoubtedly, data quality plays a key role. As Grellety and Golden (2016) showed, higher random measurement errors lead to larger standard deviations [5]. In other words, large deviations not only inflate estimates of stunting, wasting, and underweight, but also overestimate over-tallness and overweight/obesity [5]. DHS data quality reviews suggest that, while heterogeneous populations might theoretically show wider SDs, there are case studies that demonstrate how procedural rigor can overcome demographic complexity. An example is Colombia’s strict adherence to WHO guidelines, likely responsible for its high-quality survey data [42].

Yet there is equally compelling evidence for high implausible values being a marker of higher social inequities [26]. The flagging of cut-offs has been challenged based on observations of living children whose z-scores are beyond currently defined implausible values [43].

In our sub-national analysis from India, we observed a decreasing trend in implausible values and standard deviations of z-scores, which could be a product of both reductions in inequities and improvements in data quality procedures. However, no state or survey reported implausible values that could meet the cut-off of 1%, thus raising further questions on the flagging of cut-offs.

### Limitations

While our analysis considered demographic representativeness for sex and age, it did not comprehensively address residence in rural or urban areas, which may be inaccurately accounted for in the latest surveys. We did not propose any post-hoc corrections for random errors, prioritizing instead procedures to prevent future errors through enhanced training and protocols [16, 44]. Finally, assessing the implementation quality of data quality procedures via direct observation and/or qualitative study was beyond this study’s scope.

### Policy and program implications

This study underscores the critical impact of data quality procedures on national and state-level decision-making. Poor data quality can obscure progress against malnutrition, lead to misallocation of resources, misguide policymakers’ efforts, and weaken confidence in crucial nutrition programs. Enhancing data quality requires continuous improvements in survey planning, training, data collection, real-time monitoring, analysis, and reporting [45, 46]. Centralised tracking of data collector abilities may strengthen baseline personnel capabilities [50].

Our findings also show a synergistic relationship between administrative and survey data [47–49]. Well-maintained administrative data bolsters confidence in survey data by enabling cross-verification [49]. Good administrative data is also likely to be accompanied by good survey data, as both are outgrowths of a robust overall public health and data ecosystem, and both are foundational to effective childhood nutrition policy [51,52].

In summary, enhanced survey methodologies in India at the national and state level have improved anthropometric data quality, but further improvement is still needed. However, not every issue is a sign of poor data quality. We have observed that the consistently high percentage of flagged values, despite tight SDs, suggests inequitable health outcomes.

We recommend prioritizing synchronized methodologies for consistent temporal comparisons, strengthened survey planning, training, and robust real-time monitoring, with transparent reporting of data quality metrics. We also suggest caution in interpreting implausible values solely as a data quality issue. Robust administrative data systems, complemented by rigorous survey data, are crucial for data quality. Transparency across all data sources, whether administrative or from surveys, is essential for both trust and data quality.

## Data Availability

The National Family Health Survey (NFHS) data can be accessed through the Demographic and Health Surveys (DHS) Program website at https://www.dhsprogram.com/. The Comprehensive National Nutrition Survey (CNNS) data, endorsed by the Ministry of Health and Family Welfare (MoHFW), Government of India, are available to researchers upon submission of a valid request to the appropriate authorities.

https://www.nfhsiips.in/nfhsuser/datarequest.php

## Ethics approval

This study involved secondary data analysis of existing datasets. Ethical clearance for the study was obtained from the Institutional Ethics Committee of the Postgraduate Institute of Medical Education and Research, Chandigarh.

## Acknowledgements

We extend our heartfelt gratitude to the late Dr. M.K. Bhan for his exemplary leadership in advancing evidence-based decision-making through the Knowledge Integration and Translation (KnIT) platform, supported by Grand Challenges India at BIRAC, a collaboration between Department of Biotechnology and Gates Foundation. We further extend thanks to Gates Ventures for their ongoing support of global health research initiatives. We also express our special thanks to the Technical Advisory Group and the KnIT Scientific and Advisory Group. Deep appreciation goes to Dr. Shirshendu Mukherjee, Dr. Chandra Madhavi, and Dr. Debanjana Dey for their technical expertise, and to Dr. Karin Lapping, Mr Ryan Fitzgerald, and Mr. Jamal Yearwood from Exemplars in Global Health Program for their invaluable collaboration. We acknowledge the unwavering support of Dr. Podugu Sainath, for the successful completion of this work. Finally, we thank Mr. David W. Stoesz for copyediting support.

## Author Contributors

**Conceptualization:** Robert Johnston, Phuong H. Nguyen, Purnima Menon, Rajesh Kumar, Drishti Sharma.

**Methodology:** Robert Johnston, Phuong H. Nguyen, Purnima Menon, Rajesh Kumar, Sarang Pedgaonkar, S.K. Singh, Drishti Sharma, Pandara Purayil Vijin.

**Analysis and visualisation:** Pandara Purayil Vijin, Drishti Sharma.

**Writing – original draft:** Drishti Sharma, Pandara Purayil Vijin, Robert Johnston, Sarang Pedgaonkar, Nadia Akseer, Phuong H. Nguyen

**Writing – review & editing:** Rajesh Kumar, Purnima Menon, S.K. Singh, and Zulfiqar A Bhutta.

## Patient and Public Involvement Statement

It was not appropriate or possible to involve patients or the public in the design, or conduct, or reporting, or dissemination plans of our research

